# Extended lifetime of respiratory droplets in a turbulent vapour puff and its implications on airborne disease transmission

**DOI:** 10.1101/2020.08.04.20168468

**Authors:** Kai Leong Chong, Chong Shen Ng, Naoki Hori, Rui Yang, Roberto Verzicco, Detlef Lohse

**Affiliations:** Physics of Fluids Group, Max Planck Center for Complex Fluid Dynamics, J. M. Burgers Center for Fluid Dynamics and MESA+ Research Institute, Department of Science and Technology, University of Twente, 7500AE Enschede, The Netherlands; Gran Sasso Science Institute - Viale F. Crispi, 7 67100 L’Aquila, Italy; Dipartimento di Ingegneria Industriale, University of Rome ‘Tor Vergata’, Roma 00133, Italy; Max Planck Institute for Dynamics and Self-Organisation, 37077 Göttingen, Germany; These authors contributed equally: Kai Leong Chong, Chong Shen Ng; These authors jointly supervised this work: Roberto Verzicco, Detlef Lohse

## Abstract

To mitigate the COVID-19 pandemic, it is key to slow down the spreading of the life-threatening coronavirus (SARS-CoV-2). This spreading mainly occurs through virus-laden droplets expelled at speaking, screaming, shouting, singing, coughing, sneezing, or even breathing [1–7]. To reduce infections through such respiratory droplets, authorities all over the world have introduced the so-called “2-meter distance rule” or “6-foot rule”. However, there is increasing empirical evidence, e.g. through the analysis of super-spreading events [6, 8–11], that airborne transmission of the coronavirus over much larger distances plays a major role [1–3, 7, 12–15], with tremendous implications for the risk assessment of coronavirus transmission. It is key to better and fundamentally understand the environmental ambient conditions under which airborne transmission of the coronavirus is likely to occur, in order to be able to control and adapt them. Here we employ direct numerical simulations of a typical respiratory aerosol in a turbulent jet of the respiratory event within a Lagrangian-Eulerian approach [16–18] with 5000 droplets, coupled to the ambient velocity, temperature, and humidity fields to allow for exchange of mass and heat [19] and to realistically account for the droplet evaporation under different ambient conditions. We found that for an ambient relative humidity of 50% the lifetime of the smallest droplets of our study with initial diameter of 10 µm gets extended by a factor of more than 30 as compared to what is suggested by the classical picture of Wells [20, 21], due to collective effects during droplet evaporation and the role of the respiratory humidity [22], while the larger droplets basically behave ballistically. With increasing ambient relative humidity the extension of the lifetimes of the small droplets further increases and goes up to 150 times for 90% relative humidity, implying more than two meters advection range of the respiratory droplets within one second. Smaller droplets live even longer and travel further. Our results may explain why COVID-19 superspreading events can occur for large ambient relative humidity such as in cooled-down meat-processing plants [10] or in pubs with poor ventilation. We anticipate our tool and approach to be starting points for larger parameter studies and for optimizing ventilation and indoor humidity controlling concepts, which in the upcoming autumn and winter both will be key in mitigating the COVID-19 pandemic.

---

Airborne transmission of COVID-19 by respiratory aerosols consisting of small and tiny saliva and mucus droplets is increasingly believed to be a crucial factor in the spreading of the pandemic, in particular in indoor situations [1–3, 7, 12–15]. Hitherto laboratory studies have focused on the virological side, by investigating the viral load of the droplets [23]. Unfortunately, surprisingly little is known on the actual fate of the respiratory droplets, once they have been expelled. Such knowledge, however, is vital to reduce the number of infections and the reproduction factor R of COVID-19. Many key questions are intimately related to fluid dynamics and flow physics: (i) How many droplets are actually expelled at the different respiratory events? (ii) What is their initial size distribution? (iii) What is the lifetime of these respiratory droplets and how does it depend on humidity and temperature? (iv) To what degree do collective effects inside the humid cloud of droplets play a role? (v) How do the aerosol droplets distribute, in particular indoors, and what are ventilation concepts to get rid of them?

The answers to all of these questions are key to reduce the further spreading of COVID-19 [6, 7, 24]. Up to now, without sufficient answers to above questions, the authorities attempt to reduce the spreading with social distancing and the so-called ‘distance rule’, which in fact had already been suggested by G. A. Soper in his 1919 Science article “The lessons of the pandemic” (then Spanish Flue): The distance between people should not be less than one and a half, or better yet, two meters (“six-foot rule”). This rule is based on a theory of viral infection by droplets from the 1930s and earlier. The picture that William F. Wells developed at that time [20, 21] in connection with the transmission of tuberculosis was the following: The drops produced by sneezing and coughing would have a wide size distribution and would fly out of the mouth and nose without much interaction between them. The small droplets would hardly be a problem because they would evaporate very quickly in the air and leave dry and therefore – as was thought – less dangerous aerosol particles behind, while the large droplets would behave ballistically. In this model, which is still used for risk assessment, the border between large and small is set at a droplet diameter of d = 5 - 10 µm. When the droplets have a diameter larger than 5 to 10 µm, the World Health Organisation (WHO) defines them as respiratory droplets and the corresponding infections as host-to-host route, while smaller droplets are referred to as droplet nuclei and the infections as aerosol route [25, 26]. For comparison: air drag on droplets becomes negligible beyond a diameter of *d* ≳ 100 µm and a coronavirus has a typical diameter of about 120 nm.

However, in recent months the empirical evidence that the six-foot rule is not sufficient to protectN against infection with the coronavirus has kept on accumulating and various so-called super-spreading events have been reported, see e.g. [6, 8–11, 27–29], all of them indoors. These super-spreading events suggest airborne transmission of the coronavirus [1–3, 12–14], with tremendous implications for the risk assessment of coronavirus transmission. Indeed, over the last years L. Bourouiba and coworkers have shown [1, 22, 30, 31] that the range and the lifetime of the cloud of tiny saliva and mucus droplets (called respiratory droplets or aerosols in this paper) is much larger than what the 6-foot rule assumes, namely up to 8 meters and up to 10 minutes, instead of one to two meters and less than one second. The reason for this is that the droplets of saliva and mucus are expelled together with warm and humid air, which considerably delays their evaporation. In addition, they are ejected as a cloud, whereby they protect each other against evaporation, so to speak: The rate of evaporation is determined by the moisture gradient on the surface of the droplets. This is, of course, much smaller in a cloud of droplets where each of the droplets releases water vapor to the environment than for individual, isolated droplets. These two effects together can easily considerably delay the evaporation of the small aerosol droplets [1]. Indeed, as Villermaux and coworkers pointed out [32, 33], the lifetime of droplets in an aerosol is determined by the turbulent mixing process, rather than by the so-called “*d*^2^-law”, which only applies to an *isolated* evaporating spherical droplet, whose square of its diameter *d* linearly decreases with time [34] and on which the estimates by Wells [20, 21] were based. The vastly different fate of droplets in a humid cloud of other droplets have also been demonstrated for evaporating dense sprays [32, 33] and for the vaporization of combustion fuels [35, 36].

An increasing number of studies – both empirical, from the medical side, from fluid dynamics, and from the aerosol side – is supporting the view that long-distance airborne transmission through multiphase turbulent droplet cloud emission is an essential factor [1–7, 12, 24, 37–41]. Prather *et al*. [6] give strong evidence that SARS-CoV-2 is spreading over large distances in aerosols exhaled by highly contagious infected individuals with no symptoms, a view supported in refs. [13, 15, 42, 43]. Owing to their small size, aerosols may lead to higher severity of COVID-19 because virus-containing aerosols penetrate more deeply into the lungs. These aerosols can accumulate, remain infectious in indoor air for hours, and be easily inhaled deep into the lungs.

Interestingly, in spite of the major research effort of the last months in particular, the role of environmental factors such as temperature and humidity remains controversial and inconclusive [44, 45]. Various researchers have tried to establish empirical relations between the temperature and the humidity and the number of infections, but given that presumably most infections have happened indoors, it is not surprising that such efforts have remained unsuccessful [7, 44, 45].

The main difficulty in getting conclusive and reproducible results originates from the lack of controlled conditions under which the spreading events occur. However, controlled experiments under well-defined conditions such as flow rate, droplet size distribution, temperature, and relative humidity are very difficult to achieve and even with controlled and reproducible conditions, following 1000s of microdroplets in space and time remains extremely challenging.

A good alternative thereof are direct numerical simulations. These not only allow to follow 1000s of microdroplets in turbulent flow, but also to straightforwardly vary the control parameters such as the droplet density, the type and strength of ventilation, the ambient temperature and the ambient relative humidity, in order to elucidate how these control parameters affect the lifetime of the respiratory droplets. In this way we want to elucidate the flow physics of these droplets, with the ultimate goal to be able to take suitable countermeasures against the spread of the coronavirus and thus to reduce the COVID-19 reproduction factor R.

However, such direct numerical simulations are highly nontrivial, too. It has been attempted with Euler-Lagrangian approaches with Reynolds-averaged Navier-Stokes (RANS) approximations for a single droplet [46], for multiple droplets [47–49] and large eddy simulations (LES) [50], which however are insufficient to properly resolve the small scales of the mixing process, which are crucial for the droplet evaporation. Also temperature and humidity fields are not fully coupled, which however determine the evaporation rate and thus the lifetime of the droplets and are essential to properly describe their collective effects. The importance of these couplings have indeed been recognized in simulations of other aerosols, such as in combustion [51]. Other attempts include simulations with continuum models [52, 53] or mathematical models [54, 55].

To overcome these difficulties and to take the full physics into account, here we perform direct numerical simulations of a respiratory event *without flow modelling*, with the temperature and humidity field fully coupled to the Navier-Stokes equation, and 1000s of droplets of a given initial size distribution (adopted from experiment [56]) treated in an Euler-Lagrangian way, with mass and temperature exchange with the environment to allow for evaporation and condensation, similarly as ref. [57]. This is only possible thanks to a highly efficient and parallelized advanced finite difference Navier-Stokes solver (“AFiD”) based on a second-order finite difference scheme [58], coupled to the advection equations for temperature and vapor concentration, both in Boussinesq approximation [59]. Details of the numerical scheme and setup are given in the Method Section (supplementary material). The objective of this numerical work is not only to elucidate the flow physics of a respiratory event and in particular how and why the lifetime of the respiratory droplets is hugely increased as compared to the case of isolated droplets and how this depends on the ambient conditions such as the ambient humidity, but also to sketch the essential ingredients of a numerical tool, which can be used for further parameter studies and more complex situations of respiratory events, in particular in indoor situations.

## Numerically simulating a respiratory event

We simulated a turbulent puff sustained over a duration of 0.6 s into ambient air and laden with 5000 water droplets, mimicking a strong cough. In addition to the droplets, the turbulent puff expels hot, vapor saturated air with an initial temperature 34 °C and relative humidity 100% [22]. Both temperature and vapor fields are buoyant. As reference ambient conditions, we chose the typical indoor ambient temperature of 20 °C and varying relative humidity between 50% and 90%. The background airflow conditions can also be an important factor, e.g. with or without ventilation [56]. Here we chose a quiescent background field, but different types of ventilation could be embodied straightforwardly.

The topic of distribution of the initial droplet sizes is in itself a subject of considerable importance and active debate [56,60,61]. Here, we seeded the respiratory event with droplets with initial diameters ranging between 10 µm and roughly 1000 µm, based on an experimental measurement [56]. Note that even smaller droplets could be added, but they would further extend the required CPU time and are not necessary to convey the main message of this work. The droplets and the turbulent puff are two-way coupled [57], that is, the droplets exchange heat and vapor mass to their surroundings. Given the dilute nature of the dispersed phase, here, we neglected the momentum exchange. Post-expulsion, we tracked the cough and droplets up to several seconds, which is sufficient to reveal the physics of their collective evaporation.

For more details on the underlying equations, the numerical simulations, and the values of the control parameters, both dimensional and non-dimensional, we refer to the Methods Section.

## Considerably extended lifetime of the smaller droplets

We first describe our results for fixed ambient relative humidity of RH = 50%. Within about 200 ms from the start of the cough, droplets larger than about 100 µm are observed to immediately fall out from the ‘puff’, basically behaving ballistically. The associated distances of this fallouts typically range between 0.1 m and 0.7 m from the source (Fig.1d). Indeed, this type of fallout has already been predicted in the 1930s by Wells [20, 21], and also demonstrated in the cough and sneeze experiments by [22]. These typical distances appear to be the basis of the spatial separation guidelines issued by the WHO, CDC, and European Centre for Disease Prevention and Control on respiratory protection for COVID-19 [15].

**Figure 1:**
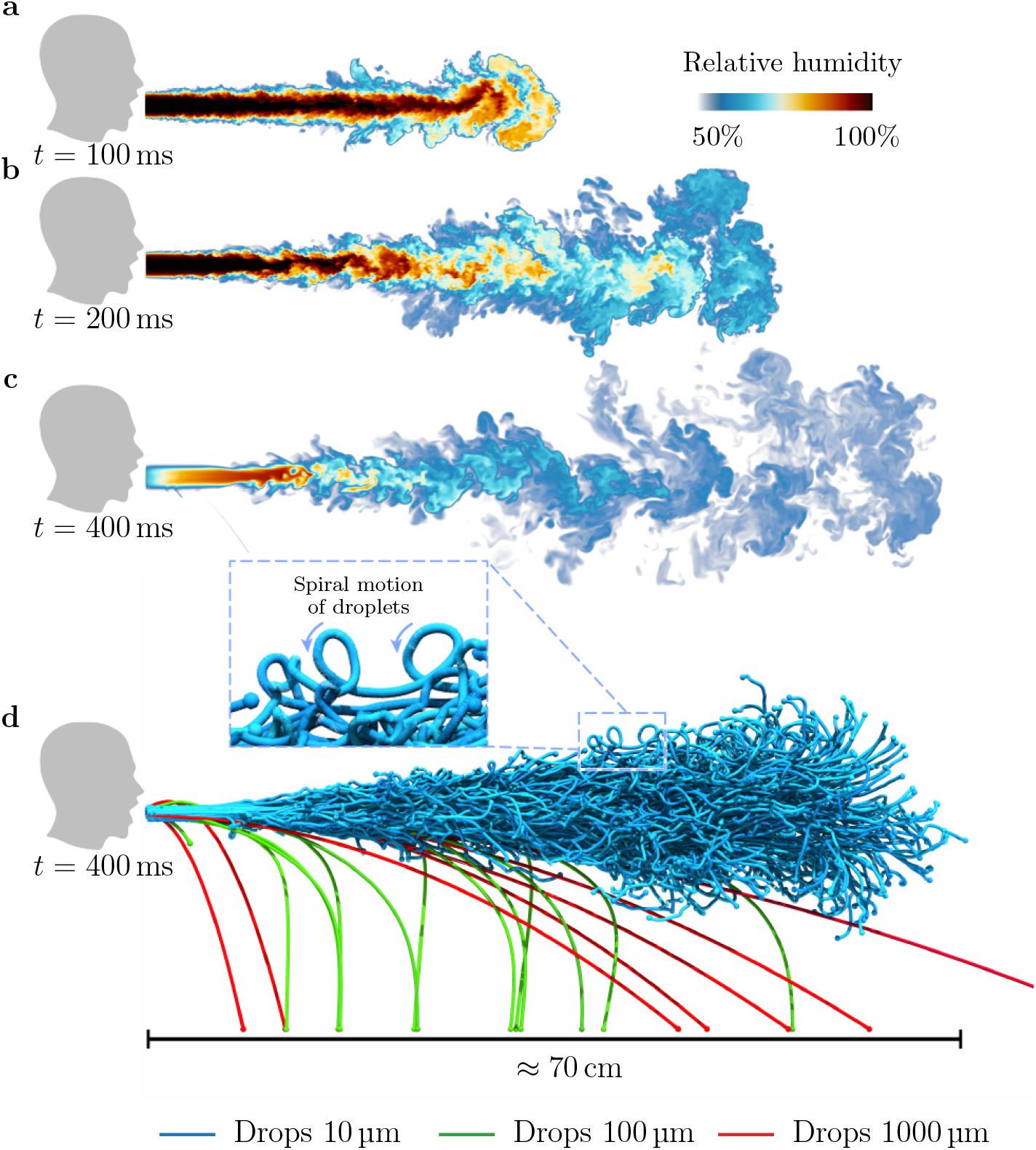
Visualisations of droplets in a heavy cough for RH = 50%: **a-d**, Snapshots of the droplet-laden cough simulation. At time *t* = 100 ms, the cough contains hot air with high moisture content. The hot moist air propagates (*t* ≈ 200 ms) and dissipates (*t* ≈ 400 ms) into the ambient surroundings. At *t* ≈ 400 ms, we show larger droplets falling out from the puff whereas smaller droplets remain protected and are carried along by the puff.

However, droplets smaller than 100 µm behave completely differently. Note again that, according to the WHO definition, respiratory infections transmitted by these smaller droplets (diameter *d* ≤ 100 µm) are considered to be occurring through the droplet transmission route. Here we see that they are airborne anyhow: Indeed, whilst larger droplets basically fall downwards like a projectile following a parabola, the paths traced by such smaller droplets remain largely horizontal and form spirals, consistent with the airborne infection scenario.

The physical explanation of the two very different behaviors is straightforward: Larger droplets fall under the influence of their own weight and are unperturbed by the surrounding airflow. In contrast, smaller droplets settle slower than the characteristic velocity of the surrounding fluid, and are therefore advected further by the local turbulent flow. This latter mechanism is intimately related to the ‘airborne’ transmission route for infectious diseases [62]. Thus, smaller droplets are clearly heavily influenced by the turbulent dispersive processes.

That the smaller droplets tend to remain inside the humid puff has dramatic consequences on their *lifetimes*, which far exceed those of isolated droplets (Fig.2a,d). Here we report results for ambient relative humidity (RH) of 50% and 90%, respectively. In the former case (RH = 50%), the droplets of 10 µm can live up to 60 times longer than the expected value by Wells, whereas in the latter case (RH = 90%), it can even become 100 to 200 times longer. These extended lifetimes are also confirmed by the much slower shrinkage rates of the droplet surface area as compared to the corresponding rate determined from the *d*^2^-law, which is valid for isolated droplets and which is the basis for Wells’ theory and the 6-foot distance rule, see Fig.2b,e.

**Figure 2:**
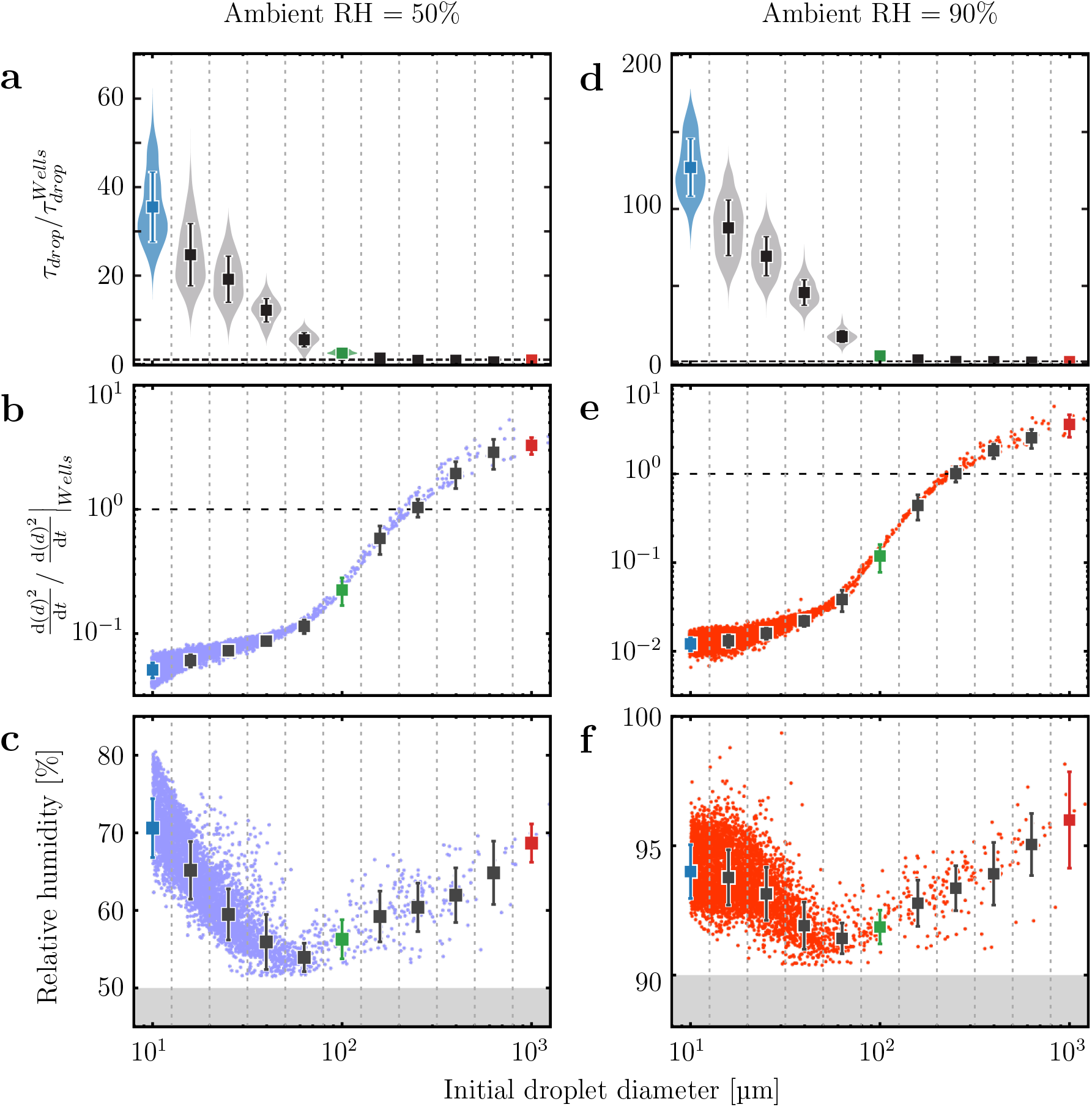
Statistics of droplets for different ambient RH. **(a-c)**, ambient RH = 50% and **(d-f)**, ambient RH = 90%. Graphs showing statistics of droplets plotted against their initial diameters, *d*. **a**,**d**, Lifetime distribution of droplets compared to lifetime estimation from Wells [20] (dashed horizontal line). **b**,**e**, Averaged change of the surface area of a droplet throughout its lifetime compared to that from Wells’ estimation (dashed horizontal line). **c**,**f**, Averaged relative humidity. For illustrative purposes, different colours denote different initial droplet diameters: blue (*d* = 10 µm), green (*d* = 100 µm) and red (*d* = 1000 µm). Plots show mean values with one standard deviation ranges.

We now further describe the flow physics contributing to this highly extended lifetime of the small droplets. The first physical factor depends on the motion of droplets relative to their surrounding fluid. As shown in Fig.1d, smaller droplets have the tendency to be captured by the turbulent puff and move together with the fluid. This gives rise to smaller relative velocities and hence less evaporation due to the reduction of convective effects. In contrast, larger droplets tend to fly and settle faster than the surrounding fluid, thus evaporating faster than predicted by the *d*^2^-law, because the convective effects, carrying the evaporated vapor away from the droplet, are considerable. This rapid evaporation is shown in the faster shrinkage rate of the droplet surface areas in Fig.2b,e.

The second and more crucial factor is the influence of the humid air around the small droplets, originating from the humid puff and the ambient surroundings. In order to quantify this effect on the lifetime, we show the averaged relative humidity of the droplets throughout the simulation as function of the initial droplet diameter *d* in Fig.2c,f. From this figure, we observe a clear non-monotonic behavior reflecting two different regimes. In the first regime for small droplets *d* = 10 − 100µm, the relative humidity takes higher values than the ambient, reflecting that the droplets are surrounded by nearly saturated humid vapor. As the initial droplet size increases, the relative humidity decreases because the settling speed increases and the droplets tend to stray from the puff (Fig.1d). In the second regime, for large droplets *d* > 100 µm, however, we observe that the relative humidity increases with the increase in size. The reason is that larger droplets evaporate larger quantities of vapor per given time, which leads to higher relative humidity in their surroundings. This effect is very local, due to the strong shear around the falling droplets.

## Propagation of the turbulent humid puff

Since the humid puff leads to extended droplet lifetimes, it is instructive to examine the propagation of the humidity field. Indeed, also in the case of dense sprays [32, 33], the droplets’ fate is determined by the vapor concentration field. Therefore, as pointed out above, the problem of droplet evaporation in a respiratory puff is intimately related to scalar mixing in turbulent flow [63, 64].

In Fig.3a,b, we show the relative humidity in the puff as function of time and distance from the respiratory release event, in order to quantify the propagation of the puff after exhalation. Very soon after the respiratory event starts, moist air coming out of the mouth creates very high humidity region in the vicinity ≈ 0.3 m of the mouth in 0.2 s. Although the overall humidity rapidly decreases after the respiratory event stops, the puff continues to propagate because of the conservation of momentum with the puff edge growing proportionally as *t*^1/4^ as predicted by Bourouiba *et al*. [22], see the solid line in Fig.3, which is an upper bound to our numerical results.

**Figure 3:**
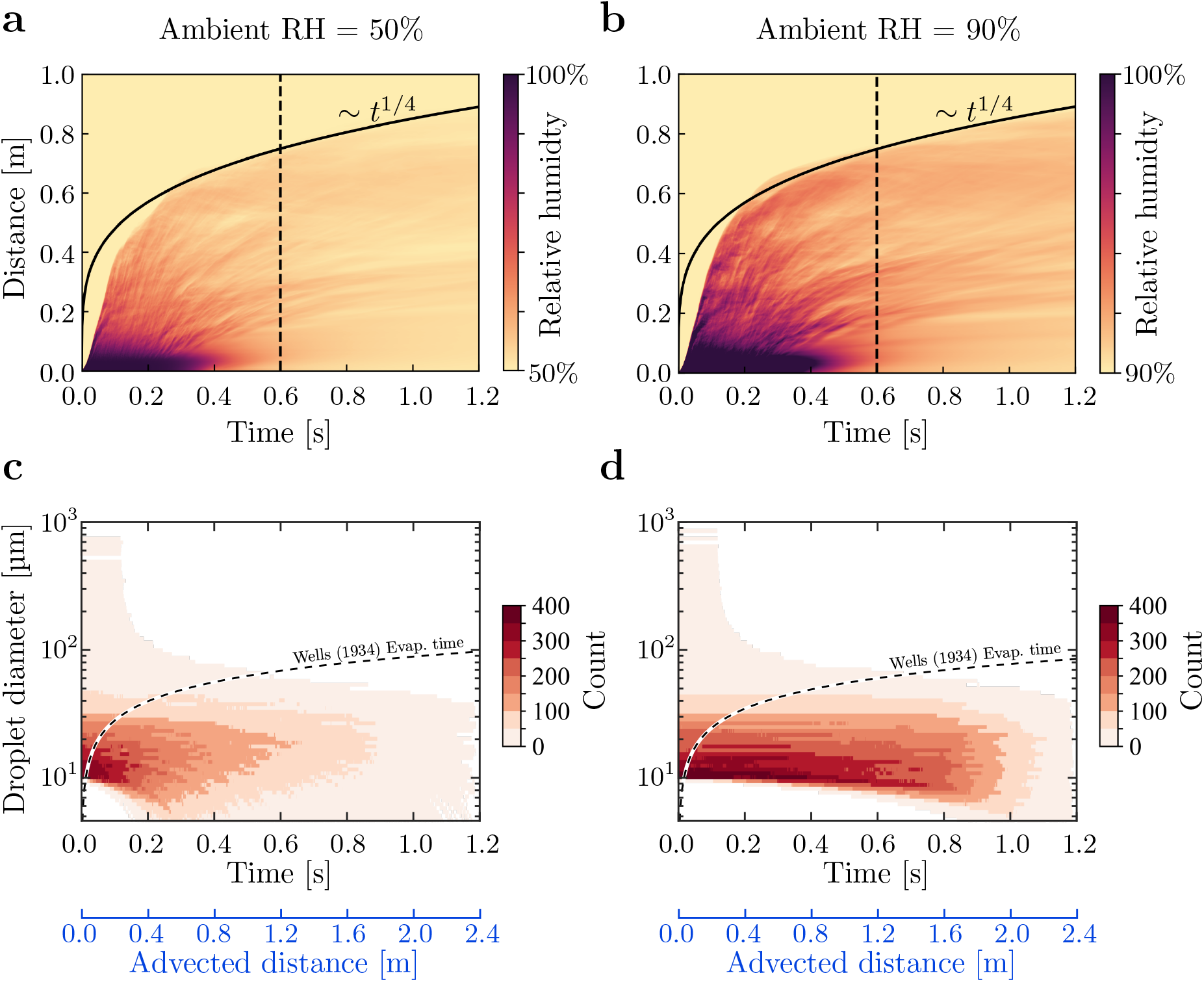
Spatial-temporal variation of relative humidity for ambient RH = 50% and 90%: **a**,**b** Relative humidity variation in space and time. The dark violet colour indicates the region with high relative humidity which can protect the droplets from shrinking. The vertical dashed line indicates the moment at which the coughing stops. After the stoppage of the coughing, the distance of the puff has the power-law relationship with time with scaling exponent 1/4, which can be obtained based on the conservation of momentum. **Comparison of expected droplet lifetime for different droplet sizes and for ambient RH = 50% and 90%: c**,**d** Count histogram of droplets at a given size and time. Time is shifted to the expulsion time for each droplet. The dashed line delineates the expected droplet lifetimes that completely evaporate, which is computed according to the assumptions by Wells [20], i.e., based on the *d*^2^-law. According to this assumption, droplets should only exist above the dashed line, which, as can clearly be seen here, is not the case in the actual respiratory event. The blue axes below panels **c** and **d** show estimated advected distance of small droplets based on a cycle-averaged cough velocity of 2 m s^−1^. Even smaller droplets with initial diameter below 10 µm live even longer and are advected even further.

Here we have considered a heavy, but single cough scenario, having 0.6 s duration. It should be noted that the travel distance and speed can be much further and faster in the case of continuous talking or successive coughing or sneezing [1, 50]. The question on how far the puff can spread in these cases can be answered based on our single cough simulation. The spatial-temporal plot in Fig.3a,b show that the edge of the puff propagates much faster than *t*^1/4^ before the end of coughing. Therefore, for successive coughing with the puff advecting at a cycle average speed of 2 - 3 m/s, the edge of puff can easily exceed 2 m in a second, while most of the droplets around 10 µm are still not evaporated because of the extended lifetime favored by the vapour puff. Droplets with an initial diameter even smaller than 10 *µ*m will travel even further and live even longer.

## Evaporation-falling curves and histograms

The extended lifetime of the small droplets can also be expressed in the so-called evaporation-falling curve, as introduced in the classical work by William F. Wells [20]. He derived the dependence of the lifetime of the droplet on its size, based on the *d*^2^-law for evaporation of an isolated droplet and on the droplet settling time, see the dashed horizontal curves in Fig.3c,d. According to this classical theory by Wells, droplets below this line evaporate completely and thus should not exist. In those figures we now include the histograms of the counts of droplets at given size and time, based on our direct numerical simulations of the respiratory events at an RH of 50% and 90%, respectively. Note that the time (on the *x*-axis) is shifted to the expulsion time for each droplet, such that the history of each droplet begins at time = 0 s. Fig.3c,d clearly show that a considerable number of small droplets do exist below the classical Wells curve, demonstrating that the classical Wells estimate is inappropriate and that small droplets can live much longer. If a background flow exists or in the case of successive respiratory events, these long-lived droplets can easily travel much farther than 2 m in a second as carried by the turbulent puff, as shown in the advected distances estimated in Fig.3c,d.

## Dependence on the ambient relative humidity RH

Up to now we have focused on two cases, namely with ambient relative humidity of 50% and of 90%, respectively. Now we repeat the numerical simulations for further ambient relative humidities in the window 50% RH 90%. From Fig.4a one can observe that the lifetime of the small droplets increases dramatically and even diverges to infinity at RH = 100%. The smaller the droplets, the more pronounced the effect. For the smallest respiratory droplets of this study with initial diameter *d* = 10 µm, for RH = 90% the lifetime extension is with a factor of about 127 as compared to the lifetime of a droplet behaving according to the Wells model, and even with a factor of 166 as compared to the lifetime of Wells model droplet at RH = 50%. Similarly, for slightly larger droplets with *d* = 20 µm, the lifetime extension remains significant with a factor of 84 and 109, respectively. The first reason for the dramatic increases in droplet lifetime seen in Fig.4a is a significantly reduced evaporation rate for larger ambient RH as the ambient gas is much closer to the saturated condition. What however also considerably contributes is that for larger ambient RH the vapour puff can be sustained for longer times and distances, as shown in Fig.4b. In such case, there is stronger protection from the vapour puff for larger ambient RH.

**Figure 4:**
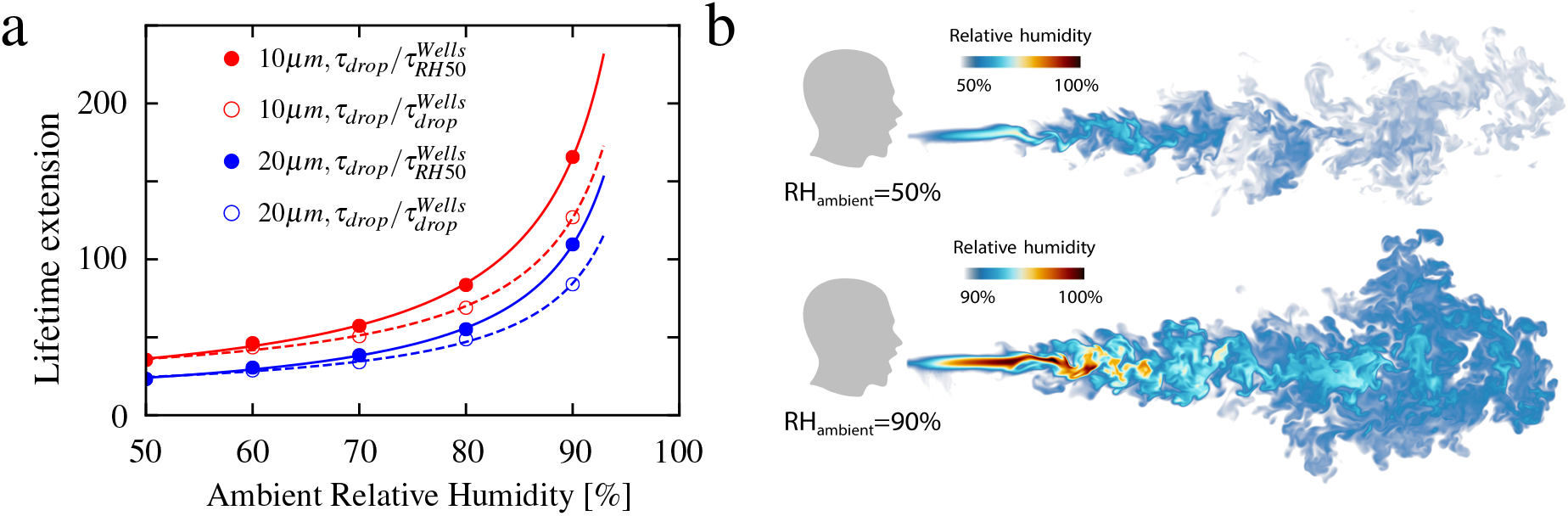
Lifetime ratio for 10 µm and 20 µm droplets: **a**, Extended lifetime as a function of relative humidity up to RH = 90%. The curves in the figure are fitted according to the function *y* = *a*_1_/(1 − *x*) + *a*_2_, where *a*_1_ and *a*_2_ are the fitting parameters. **Visualisations of humid puff for ambient RH = 50% and 90% at time** 600 ms**: b**, The humid puff maintains coherence for longer time and at much longer distances for larger ambient RH. Note the different humidity color scales for two shown cases.

## Discussions

By tracking the interactions between respiratory droplets with the local velocity, temperature, and vapour field within direct numerical simulations, we have demonstrated the huge difference between droplet lifetimes in an actual respiratory event, as compared to that predicted according to Wells’ assumption. Our numerical results are qualitatively consistent with Bourouiba’s multiphase cloud emission model [1] and, in fact, quantify it, but are inconsistent with Wells’ classical model, on which the 6-foot distance rule is based. The reason is that Wells’ model assumes that the droplets are isolated, i.e., have no interaction with the near velocity, temperature, and in humidity fields around the droplet, which is not the case in reality. Indeed, our study has conveyed that in particular the humid vapor exhaled together with the droplets must not be neglected, as the vapor concentration around the droplet remains high during the whole respiratory event and thereafter (see Fig.2a,d), strongly contributing to the lifetime extension of the small droplets by orders of magnitude. In this sense the lifetime of the respiratory droplets is mainly controlled by the mixing [63] of the humidity field exhaled together with them, similarly as occurring for the lifetime of evaporating dense sprays, which is also controlled by the mixing of the vapor field [32, 33]. The relevant length scale for droplet evaporation is therefore not the diameter of the droplet itself (sub-millimeter), but the outer length scale of the surrounding turbulent velocity and humidity field, i.e., meters.

The lifetime of respiratory droplets has, to-date, grossly been underestimated (see Fig.3). The extension of the droplet lifetime is so extreme that the smallest droplets (*d* =10 µm) of our study barely evaporate (lifetime extension by factor 35 at ambient relative humidity RH = 50% and even by factor 166 at RH = 90%) and are transported in an aerosolised manner. This finding contradicts the ‘respiratory droplet’ classification by WHO for *d* >5-10 µm droplets, which implies that droplets of these sizes fall ballistically. From our results, there is strong evidence that even smaller droplets with initial diameter *d* ≤ 10 µm will survive even far longer in aerosolised form, because of the protection from the turbulent humid puff.

Our results also show that the lifetime extension of the respiratory droplets is the more dramatic the larger the ambient relative humidity, see Fig.4a. The reason lies in the longer lifetime of the local humidity field around the droplets for larger ambient humidity RH, consistent with the picture that the mixing of the local humidity field determines the droplet lifetime, see Fig.4b. This finding may explain why many COVID-19 superspreading events have been reported in indoor environments with large ambient relative humidity. Examples are densely packed pubs with poor ventilation or meat-processing plants [10], in which the cooling down of the ambient air leads to very high indoor relative humidities. Note that high outdoor relative humidity obviously also leads to enhanced droplet lifetime, but in the context of the COVID-19 spread seems less relevant to us as most infections seem to happen indoors, due to in general sufficient ventilation outside.

To-date, the general measures to reduce the transmission of COVID-19 have mainly taken into account the mode of droplet transmission, leading to social distancing guidelines as the 6-foot rule [15]. The analysis of various super-spreading events [6, 8–11] has revealed that this is not sufficient, and our findings explain why, namely because of the dramatically enhanced lifetime of respiratory aerosols and droplets: Even droplets with a diameter of 10 *µ*m, which according to WHO terminology are called respiratory droplets and are claimed to be relevant only through the host-to-host infection route, can have a lifetime extension by a factor of 200.

Consequently, in addition to the 6-foot rule, various measures should be taken to reduce the respiratory aerosol and droplet concentration in indoor environments [1–3,6,7,12–15,24,43]. This is of particular importance in the upcoming autumn and winter when people are forced to be indoors. Soper [65], in his 1919 Science article “Lessons of the pandemic” (those days the so-called Spanish influenza) was right with his claim that “there is danger in the air in which they cough and sneeze” [65]. (With the knowledge of today, we must also add “speak”, “sing”, “scream”, and even “breath” [1, 2, 4, 13, 56].) Those days Soper came up with empirical rules to protect the population against this danger, namely (among others) “Open the windows - at home and at office” and “Suspects should wear masks” [65].

Today, we understand why Soper’s empirical rules work and are so crucial for the mitigation strategy against COVID-19 spreading. Following these rules contributes to dilute and reduce the concentration of respiratory droplets and aerosols in indoor situations [6, 24]. Our present work contributes to this understanding and our approach – direct numerical simulations with fully coupled velocity, vapor concentration and temperature fields and point-like droplets – even has the potential to furtherN quantitatively contribute:

- First, indeed, *face masks* reduce or even block respiratory droplets entering indoor environments and have turned out to be a helpful measure against the spread of COVID-19 [15, 66]. This reduction of the droplet input in indoor environments is the main function of face masks; in addition, depending on their quality and specification, they can also reduce the inhalation of respiratory droplets, which is of course essential for health-care workers dealing with COVID-19 patients.
- Second, *efficient indoor ventilation concepts* [56, 67, 68] are equally crucial, to advect the humid cloud with respiratory droplets out of the room or at least dilute its concentration. However, note that, on the one hand, the humid vapor field can be diluted and rapidly “dissipated” by ventilation, but on the other hand, it may lead to strong background flow, which results in long propagation distance. The relative strength of the two effects must be examined in detail in future research, and such indoor measures are becoming increasingly important after gradually resuming from lockdown.

Our present work, however, also suggests a further and supplementary mitigation strategy against COVID-19 infections, or, to be more precise, against high indoor respiratory droplets concentration, namely:

- *Reduction of the ambient indoor relative humidity*, as this helps to reduce the lifetime of the respiratory droplets and aerosols, see Fig.4.

Our results help to understand why these various mitigation strategies against COVID-19 are successful and legitimate, but we also anticipate that our present tool and approach will be a starting point for larger parameter studies and for further optimizing mitigation strategies such as ventilation and indoor humidity controlling concepts.

## Methods

### Governing equations

The governing equations include the equations for gas phase and equations for droplets [57]. Both temperature and vapour concentrations are coupled to the velocity field by employing the Boussinesq approximation. The motion of the gas phase is assumed to be incompressible *∂u*_*i*_/*∂x*_*i*_ = 0, and governed by the momentum, energy, and mass fraction equations:

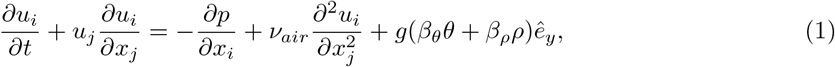

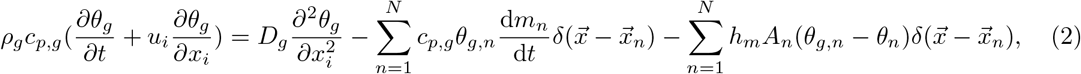

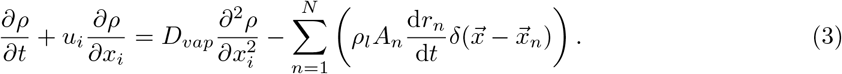

The last term on the right-hand side of equation (1) represents the coupling of temperature and mass fraction to the momentum equation.

For droplets, the spherical point-particle model is applied, and we consider the conservation of momentum (Maxey-Riley equation [69]), energy, and mass as follows:

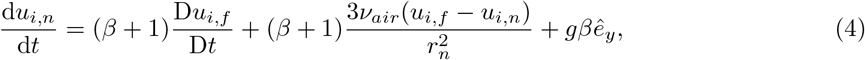

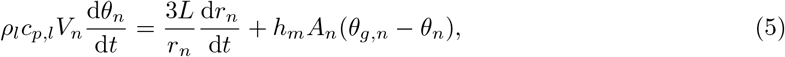

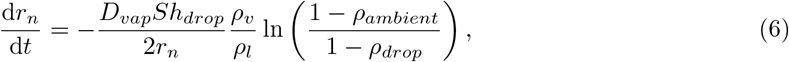

where, *u*_*i*_, *u*_*i,n*_, and *u*_*i,f*_ are the velocity of gas, droplets, and gas at the location of droplets, respectively. Similarly *θ*_*g*_, *θ*_*n*_, and *θ*_*g,n*_ are in Kelvin and used to represent the temperature of gas, droplets, and gas at the location of droplets, respectively. *ρ* is the vapour mass fraction, *r*_*n*_ the droplet radius, *A*_*n*_the surface area of the droplets, *V*_*n*_ the volume of the droplets. Also *p* denotes the reduced pressure. *h*_*m*_ is the heat transfer coefficient. *L* is the latent heat of vaporisation of the liquid.

Equation (5) and equation (6) are closed using the Ranz–Marshall correlations [70], which give us the estimation of *h*_*m*_ and *Sh*_*drop*_ for a single spherical droplet.

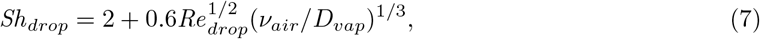

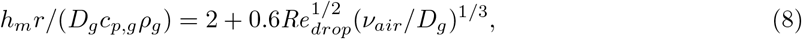

where we have droplet Reynolds number

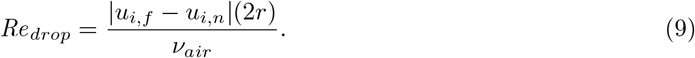

Note that, by definition, *Sh*_*drop*_ and *Re*_*drop*_ are also functions of *r*. The realistic parameters needed are listed in Extended Data Table (1) and are obtained from [22].

The relative humidity at the location of the droplets is defined as *P*_*vap,drop*_/*P*_*sat*_ = *ρ*_*vap*_/*ρ*_*sat,vap*_, where *ρ*_*vap*_ is solved from equation (3). Here, we assume that the temperature is the same. Therefore, to calculate the local relative humidity, *ρ*_*sat,vap*_ is first determined by the ideal gas law:

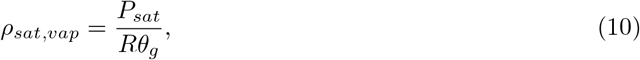

where *ρ*_*sat,vap*_, *P*_*sat*_ and *R* are the saturated vapour density, saturated pressure, and specific gas constant. And through the Antoine’s relation, the saturated pressure can be written as

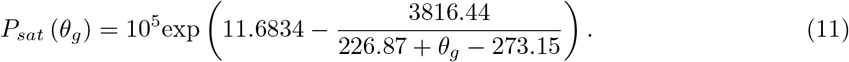

### Numerical setup

To numerically solve the equations, we used our finite difference solver AFiD [58] with high-performance Message Passing Interface (MPI) and point-particle model. The size of the computational domain in dimensional form is 0.18 m (spanwise length) ×0.37 m (height) ×1.47 m (streamwise length) and is tested to be large enough to capture the cough vapour and spreading droplets. The grid points chosen is 256 × 512 × 2048 to ensure that enough resolution has been employed.

### Cough properties

The cough profile we apply is the gamma distribution of the form *Ũ*_*cough*_(*t*) = *U*_*cough*_*αt* exp(−*αt*/4), where *α* = 60.9 s^−1^ such that the entire cough process lasts for about 0.6 s. For the initial droplet size, we employ a similar distribution as [22, 60] with 5000 droplets. The droplets are randomly positioned at the inlet and evenly injected in time.

### Lifetime statistics

For the small droplets, we compute the 80% life time, i.e. the time needed to shrink to 80% of the initial diameter. However, the large droplets rapidly settle to the boundary before reaching this threshold, so their 80% life time cannot be directly determined. To overcome this, the life times of these cases are estimated using higher diameter thresholds (see Extended Data Table (2)). The thresholds are defined such that the number of droplets satisfying the thresholds are at least 97% of the initial number. Then, we measure the shrinkage time for each droplets and calculate the ratio of the shrinkage time and to the corresponding shrinkage time with Wells’ assumption.

### Wells’ assumption

In the assumption of Wells’ model, the droplets are assumed to be unaffected by flow, so we have equation (7) and equation (8) equalling 2 and equation (5) becomes d*θ*_*d*_/d*t* = 0. The ambient temperature and ambient relative humidity remain unchanged with time. The Wells’ shrinkage rate is computed using equation (6) as

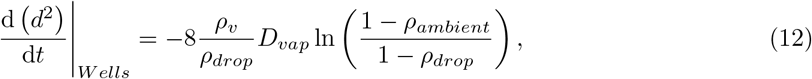

and also the lifetime as a similar manner.

## Data Availability

Additional data related to this paper may be requested from the corresponding author.

## Acknowledgements

This work was funded by the ERC Advanced Grant DDD, Number 740479 and by several NWO grants. The funders have no role in study design, data collection and analysis or decision to publish. The simulations were performed on the national e-infrastructure of SURFsara, a subsidiary of SURF cooperation, the collaborative ICT organization for Dutch education and research, and the Irish Centre for High-End Computing (ICHEC).

## Author contributions

C.S.N., K.L.C., and N.H. planned and performed code development and evaluated the data. C.S.N., K.L.C. and R.Y. performed simulations. C.S.N., K.L.C., N.H. and R.Y. analysed data. R.V. managed initial code development and supervised the simulations. D.L. initiated, designed, and supervised the project. All authors wrote the manuscript.

## Competing interests

The authors declare no competing interests.

**Correspondence and requests for materials** should be addressed to D.L.

**Extended Data Table 1:** Definition of realistic parameters employed in the numerical simulations of this study*.

| Cough and vapour properties | Symbols | Values |
| --- | --- | --- |
| The vapour mass density | $\rho_v$ | 4.85 mg/l |
| The temperature of coughing vapour | $\theta_{cough}$ | 34 °C |
| The vapour specific heat capacity | $c_{p,v}$ | 1.996 kJ/kg/K |
| The mean velocity of cough | $U_{cough}$ | 11.2 m/s |
| The diameter of mouth | $D_{mouth}$ | 2.3 cm |
| The diffusivity of water vapour in air | $D_{vap}$ | $2.5 \times 10^{-5} \text{m}^2/\text{s}$ |
| Air properties | Symbols | Values |
| The air mass density | $\rho_{ambient}$ | 1.204 kg/m<sup>3</sup> |
| The temperature of air | $\theta_a$ | 20 °C |
| The air viscosity | $\nu_{air}$ | $1.562 \times 10^{-5} \text{m}^2/\text{s}$ |
| The thermal expansion coefficient | $\beta_\theta$ | $3.5 \times 10^{-3}/\text{K}$ |
| The air specific heat capacity | $c_{p,a}$ | 1 kJ/kg/K |
| The thermal diffusivity of air | $D_g$ | $22.39 \times 10^{-6} \text{m}^2/\text{s}$ for air at 25 °C |
| Latent heat | $L$ | 2256 kJ/kg |
| Droplets properties | Symbols | Values |
| The droplet density | $\rho_l$ | 993 kg/m<sup>3</sup> |
| Density ratio term | $\beta$ | $0.5 \times 10^{-3}$ |
| The droplet specific heat capacity | $c_{p,l}$ | 4.186 kJ/kg/K |

\*Here we use the characteristic values taken from [22].

**Extended Data Table 2:** The diameter threshold and the corresponding bin range of droplet diameter.

| Threshold | Bin range of droplet diameter in logarithm ($\log(d_0)$)<br>[$\mu\text{m}$] |
| --- | --- |
| 0.8 | [1.0, 1.1] |
| 0.9 | [1.1, 1.3] |
| 0.95 | [1.3, 1.5] |
| 0.97 | [1.5, 1.7] |
| 0.99 | [1.7, 1.9] |
| 0.992 | [1.9, 2.1] |
| 0.993 | [2.1, 2.3] |
| 0.995 | [2.3, 2.5] |
| 0.999 | [2.5, 2.7] |
| 0.999 | [2.7, 2.9] |
| 0.9992 | [2.9, 3.1] |

## Notes

### Competing Interest Statement

The authors have declared no competing interest.

